# Connecting real-world digital mobility assessment to clinical outcomes for regulatory and clinical endorsement – the Mobilise-D study protocol

**DOI:** 10.1101/2022.05.25.22275598

**Authors:** A. Stefanie Mikolaizak, Lynn Rochester, Walter Maetzler, Basil Sharrack, Heleen Demeyer, Claudia Mazzà, Brian Caulfield, Judith Garcia-Aymerich, Beatrix Vereijken, Valdo Arnera, Ram Miller, Paolo Piraino, Nadir Ammour, Mark Forrest Gordon, Thierry Troosters, Alison J Yarnall, Lisa Alcock, Heiko Gaßner, Jürgen Winkler, Jochen Klucken, Christian Schlenstedt, Henrik Watz, Anne-Marie Kirsten, Ioannis Vogiatzis, Nikolaos Chynkiamis, Emily Hume, Dimitrios Megaritis, Alice Nieuwboer, Pieter Ginis, Ellen Buckley, Gavin Brittain, Giancarlo Comi, Letizia Leocani, Jorunn L. Helbostad, Lars Gunnar Johnsen, Kristin Taraldsen, Hubert Blain, Valérie Driss, Anja Frei, Milo A. Puhan, Ashley Polhemus, Magda Bosch de Basea, Elena Gimeno, Nicholas S Hopkinson, Sara C Buttery, Jeffrey M. Hausdorff, Anat Mirelman, Jordi Evers, Isabel Neatrour, David Singleton, Lars Schwickert, Clemens Becker, Carl-Philipp Jansen, members of the clinical validation study (WP4) on behalf of Mobilise-D consortium

**Affiliations:** Department of Clinical Gerontology, Robert-Bosch-Hospital, Stuttgart, Germany; Translational and Clinical Research Institute, Faculty of Medical Sciences, Newcastle University, Newcastle upon Tyne, UK; Department of Neurology, University Medical Center Schleswig-Holstein, Kiel, Germany; Department of Neuroscience and Sheffield NIHR Translational Neuroscience BRC, Sheffield Teaching Hospitals NHS Foundation Trust & University of Sheffield, Sheffield, England; Department of Rehabilitation Sciences, KU Leuven, Leuven, Belgium; Department of Respiratory Diseases, University Hospitals Leuven, Leuven, Belgium; Department of Rehabilitation Sciences, Ghent University, Ghent, Belgium; Insigneo Institute, Department of Mechanical Engineering, University of Sheffield, Sheffield, UK; Insight Centre for Data Analytics, O’Brien Science Centre, University College Dublin, Dublin, Ireland; UCD School of Public Health, Physiotherapy and Sports Science University College Dublin, Dublin, Ireland; Barcelona Institute for Global Health (ISGlobal), Barcelona, Catalonia, Spain.; Universitat Pompeu Fabra (UPF), Barcelona, Spain.; CIBER Epidemiología y Salud Pública (CIBERESP), Madrid, Spain.; Department of Neuromedicine and Movement Science, Norwegian University of Science and Technology, Trondheim, Norway; Clario, Geneva, Switzerland; Novartis Institutes for Biomedical Research, Cambridge, MA, USA; Research and Development, Pharmaceuticals, Bayer AG, Berlin, Germany; Clinical Science & Operations, Global Development, Sanofi R&D, Chilly-Mazarin, France; Specialty Clinical Development, Teva Pharmaceuticals, West Chester, PA, USA; Department of Molecular Neurology, University Hospital Erlangen, Erlangen, Germany; Digital Medicine, University of Luxembourg, Esch-sur-Alzette, Luxembourg; Institute of Interdisciplinary Exercise Science and Sports Medicine, Medical School Hamburg, Hamburg, Germany; Pulmonary Research Institute at LungClinic Grosshansdorf GmbH, Airway Research Center North (ARCN), German Center for Lung Research (DZL), Grosshansdorf, Germany; Department of Sport, Exercise and Rehabilitation, Faculty of Health and Life Sciences, Northumbria University, Newcastle, England; Thorax Research Foundation & First Dept. of Respiratory Medicine, National & Kapodistrian University of Athens, Sotiria General Chest Hospital, Athens, Greece; KU Leuven, Department of Rehabilitation Sciences, Neurorehabilitation Research Group (eNRGy), Leuven, Belgium; Department of Neurology, San Raffaele University, Milan, Italy; Casa di Cura del Policlinico, Milan, Italy; Department of Physiotherapy, Oslo Metropolitan University (OsloMet), Oslo, Norway; Department of Geriatrics, Montpellier University Hospital and MUSE, Montpellier, France; Epidemiology, Biostatistics and Prevention Institute, University of Zurich, Zurich, Switzerland; National Heart and Lung Institute, Imperial College London, London, England; Center for the Study of Movement, Cognition and Mobility, Neurological Institute, Tel Aviv Sourasky Medical Center, Tel Aviv, Israel; Department of Physical Therapy, Sackler Faculty of Medicine & Sagol School of Neuroscience, Tel Aviv University, Tel Aviv, Israel; Rush Alzheimer’s Disease Center and Department of Orthopaedic Surgery, Rush University Medical Center, Chicago, Illinois United States; McRoberts BV, The Hague, Netherlands

**Keywords:** Digital mobility outcome, performance, gait, chronic disease, Parkinson’s disease, Multiple Sclerosis, Chronic Obstructive Pulmonary Disease, Proximal femoral fracture, regulatory.

## Abstract

**Background:** The development of optimal strategies to treat impaired mobility related to ageing and chronic disease requires better ways to detect and measure it. Digital health technology, including body worn sensors, has the potential to directly and accurately capture real-world mobility. Mobilise-D consists of 34 partners from 13 countries who are working together to jointly develop and implement a digital mobility assessment solution to demonstrate that real-world digital mobility outcomes have the potential to provide a better, safer, and quicker way to assess, monitor, and predict the efficacy of new interventions on impaired mobility. The overarching objective of the study is to establish the clinical validity of digital outcomes in patient populations impacted by mobility challenges, and to support engagement with regulatory and health technology agencies towards acceptance of digital mobility assessment in regulatory and health technology assessment decisions

**Methods/Design:** The Mobilise-D clinical validation study is a longitudinal observational cohort study that will recruit 2400 participants from four clinical cohorts. The populations of the Innovative Medicine Initiative-Joint Undertaking represent neurodegenerative conditions (Parkinson’s Disease), respiratory disease (Chronic Obstructive Pulmonary Disease), neuro-inflammatory disorder (Multiple Sclerosis), fall- related injuries, osteoporosis, sarcopenia, and frailty (Proximal Femoral Fracture). In total, 17 clinical sites in ten countries will recruit participants who will be evaluated every six months over a period of two years. A wide range of core and cohort specific outcome measures will be collected, spanning patient-reported, observer-reported, and clinician-reported outcomes as well as performance-based outcomes (physical measures and cognitive/mental measures). Daily-living mobility and physical capacity will be assessed directly using a wearable device. These four clinical cohorts were chosen to obtain generalizable clinical findings, including diverse clinical, cultural, geographical, and age representation. The disease cohorts include a broad and heterogeneous range of subject characteristics with varying chronic care needs, and represent different trajectories of mobility disability.

**Discussion:** The results of Mobilise-D will provide longitudinal data on the use of digital mobility outcomes to identify, stratify, and monitor disability. This will support the development of widespread, cost- effective access to optimal clinical mobility management through personalised healthcare. Further, Mobilise-D will provide evidence-based, direct measures which can be endorsed by regulatory agencies and health technology assessment bodies to quantify the impact of disease-modifying interventions on mobility.

**Trial registration:** ISRCTN12051706

## Introduction

A key challenge for delivering healthcare in ageing societies is the optimal evaluation of mobility, which can be broadly defined as the ability and performance of a person to move about in their environment (1). Central aspects of mobility according to the World Health Organization (WHO) are changing one’s body position or location; or transferring from one place to another; carrying, moving or manipulating objects; or walking, running or climbing (2).

Walking is the most common and functionally relevant aspect of mobility that is affected by age- associated processes and multiple chronic diseases. Walking is a complex activity that requires interactions between the cardiovascular, pulmonary, and musculoskeletal systems as well as widespread brain networks for effective performance (3). Deteriorations in these systems are reflected in walking performance. As such, walking speed is increasingly denominated as the “6^th^ vital sign of health” (4) and represents an appropriate mobility measure for multiple populations. This is mirrored by strong evidence that mobility outcomes, such as walking speed and physical activity predict morbidity, mortality, falls, cognitive impairment, and disability (5–10). It is, therefore, not surprising that people living with chronic conditions often rate physical mobility – and specifically walking ability – as one of the most important clinical outcome measures (11–15).

Currently, clinical research and practice mainly rely on patient-reported outcomes (self- reported/perceived walking capacity/ability), objective clinical assessments of walking capacity, and subjective clinical assessments (clinician-led evaluation of walking capacity). All assessments are subject to recall and response bias, are often burdensome in their execution (for both patients and assessors), have ceiling or floor effects, Hawthorne effects, and/or other limitations (16–18). Fluctuations due to medication and disease exacerbation further reduce reliability and validity due to the intermittent nature of assessment. Moreover, these assessments are regularly conducted in lab-based or clinical environment which do not necessarily reflect the complex environmental determinants of functional mobility in daily life, which severely hampers their ecological validity (16, 19–21). Variations in the environment for these measurements can include supervised and controlled settings to measure capacity vs. non-supervised and uncontrolled environments (2). Despite the identified need for quantitative mobility assessment under multiple conditions, inconsistent testing procedures and wide variations in baseline “norms” have prevented the establishment of a widely adopted consensus on walking and mobility outcomes. Consequently, there is no harmonised approach to the measurement and understanding of impaired, real- life mobility.

### Rationale

There is a clinical need for mobility assessment to reflect real-world performance. In the last decade, advances in sensor technology, largely driven by the consumer market, have led to the advancement of wearable sensors that are able to record continuously for longer periods of time and include multi-sensing capabilities. Measurements can be obtained remotely (22–24), opening up opportunities to extend the scope of mobility measurement to continuously capture discrete and clinically relevant mobility characteristics. It is now feasible to conduct objective, digital mobility assessment during real-world walking, defined as unsupervised, unscripted walking behaviour which occurs in non-simulated everyday situations (16). These walking-related digital mobility outcomes (DMOs) such as volume, pace, rhythm, variability, and symmetry are increasingly used to quantify as well as qualify gait in multiple medical conditions.

Emerging evidence suggesting that DMOs are sensitive as well as ecologically valid markers of health status has evoked calls for validation and qualification by seeking regulatory approval for DMOs as clinical endpoint measures (25–30). However, studies are needed to establish their construct validity, predictive validity, responsiveness and clinical meaningfulness (31–33). The most recent systematic evidence (34, 35) supports the need for clinical validation of DMOs within Mobilise-D.

A multi-centric observational study (36) is being conducted as part of Mobilise-D to establish the technical validity and patient acceptability of the approach used to quantify digital mobility in the real-world. This technical validation study (TVS) allowed the definition of a set of procedures for the metrological verification of an inertial sensor-based device and for the experimental validation of the algorithms used to calculate the DMOs. The experimental validation included laboratory and real-world assessment in participants from five disease groups (chronic obstructive pulmonary disease (COPD), Parkinson’s disease (PD), multiple sclerosis (MS), proximal femoral fracture (PFF), and congestive heart failure). The DMOs extracted from the device were validated against those from different reference systems, chosen according to the contexts of observation. Questionnaires and interviews were used to evaluate the users’ perspective on the deployed technology and the relevance of the mobility assessment.

### Mobility constructs

Within Mobilise-D, there are three distinct mobility constructs being measured and assessed: what a person i) can do, ii) thinks they do, and iii) actually do in real life (29, 37). *Mobility capacity* represents the ability of a patient to move and is provided by supervised clinical tests such as the six-minutes walking test (6-MWT), supervised gait speed or clinician-assigned scores based on clinical tests. This is what a person <u>can do</u>. *Mobility perception* represents the patient’s subjective perception of their own mobility, and is provided through patient reported outcomes (PRO) or clinician-assigned scores based on the patient’s answers to standardised questionnaires. It captures what the patient or clinician <u>thinks the patient does</u>. *Mobility performance* represents the duration, quality, and intensity of the participant’s mobility as observed in unsupervised real-world settings, and during observational periods of time that are sufficiently long enough in duration to be considered representatives of the daily life. This is what a person <u>actually does in their real life</u>. Disparities between parameters has been shown depending on which construct was being assessed (38). There are currently no biomarkers accepted by regulators or accepted across diseases to quantify mobility performance (31–33). At present, in the regulatory evaluation of new or existing drugs targeting chronic diseases, mobility is frequently estimated in short and cohort specific lab-based assessments, rather than day-to-day performance over time (39).

### Aim and objectives

Mobilise-D Clinical Validation Study (CVS) aims to deliver and validate a new methodology for real-world digital mobility assessment to monitor and predict global and disease-specific clinical outcomes in a variety of disease states: PD, COPD, MS, and PFF. This will be used to obtain regulatory and health stakeholder approval for digital mobility assessment.

Within the clinical validation study, the objectives include:

(i) Assessing the construct validity of DMOs against established clinically relevant constructs;
(ii) Defining predictive capacity of DMOs against general and disease-specific, clinically relevant constructs;
(iii) Assessing the ability of DMOs to detect change over time in clinically relevant constructs;
(iv) Estimating the Minimal Important Difference (MID) of DMOs to measure change in disease state (worsened or improved)
(v) Describing real-world walking behaviour with DMOs in patients with PD, MS, COPD and following PFF.

## Materials and methods

The SPIRIT reporting guidelines have been followed within this manuscript (40), see Figure 1. This protocol represents version 1.5; March 2022.

**Figure 1:**
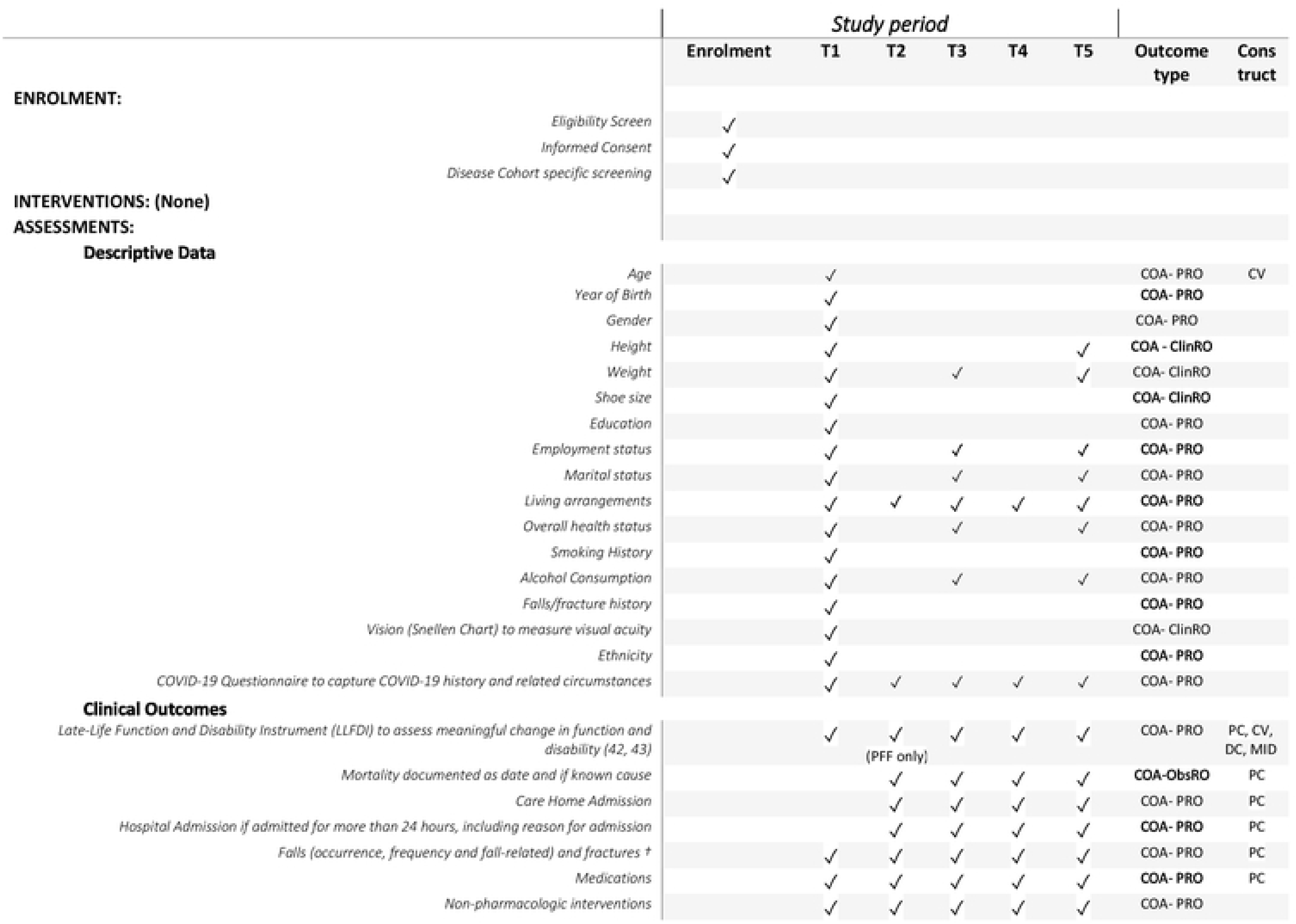
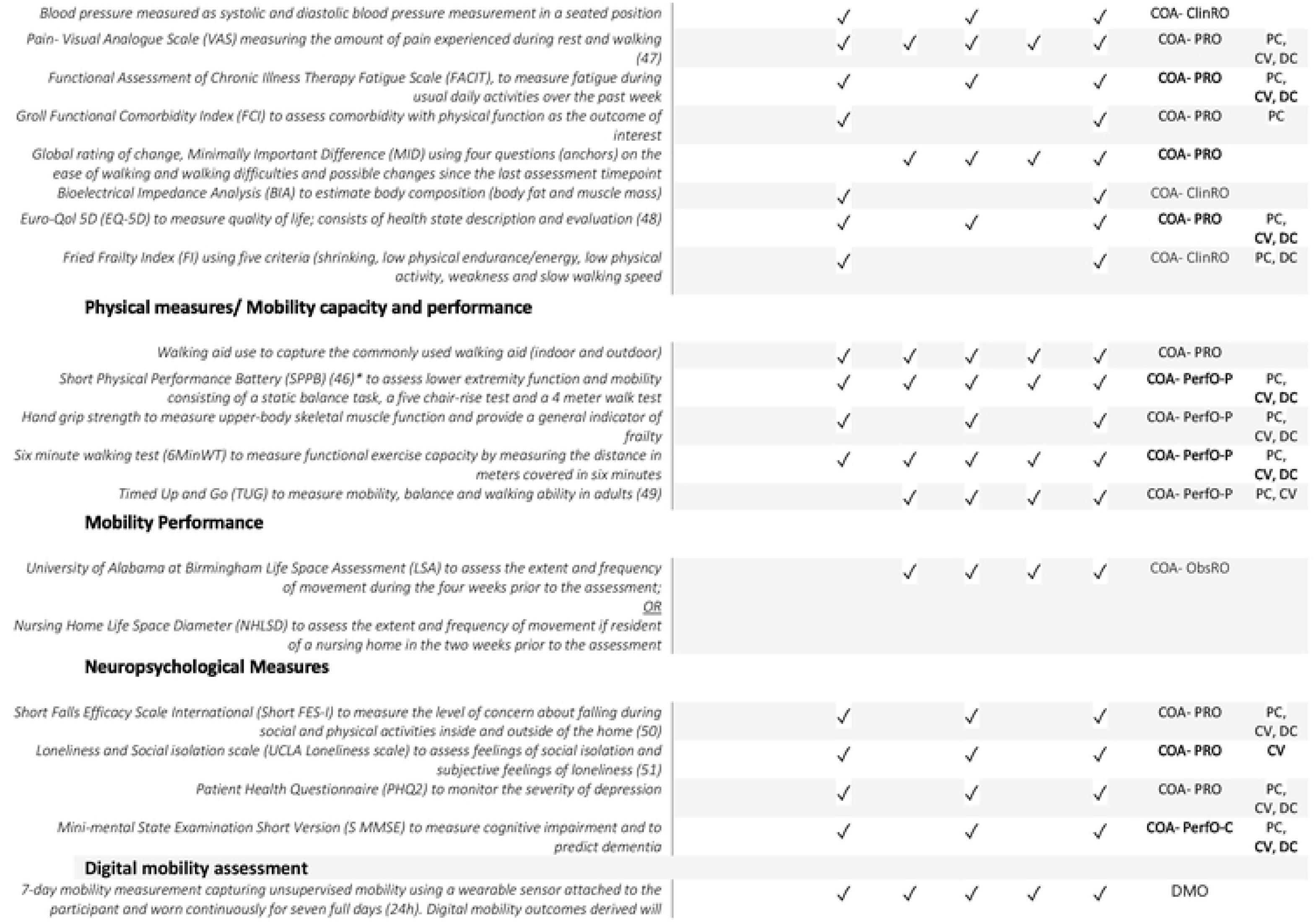
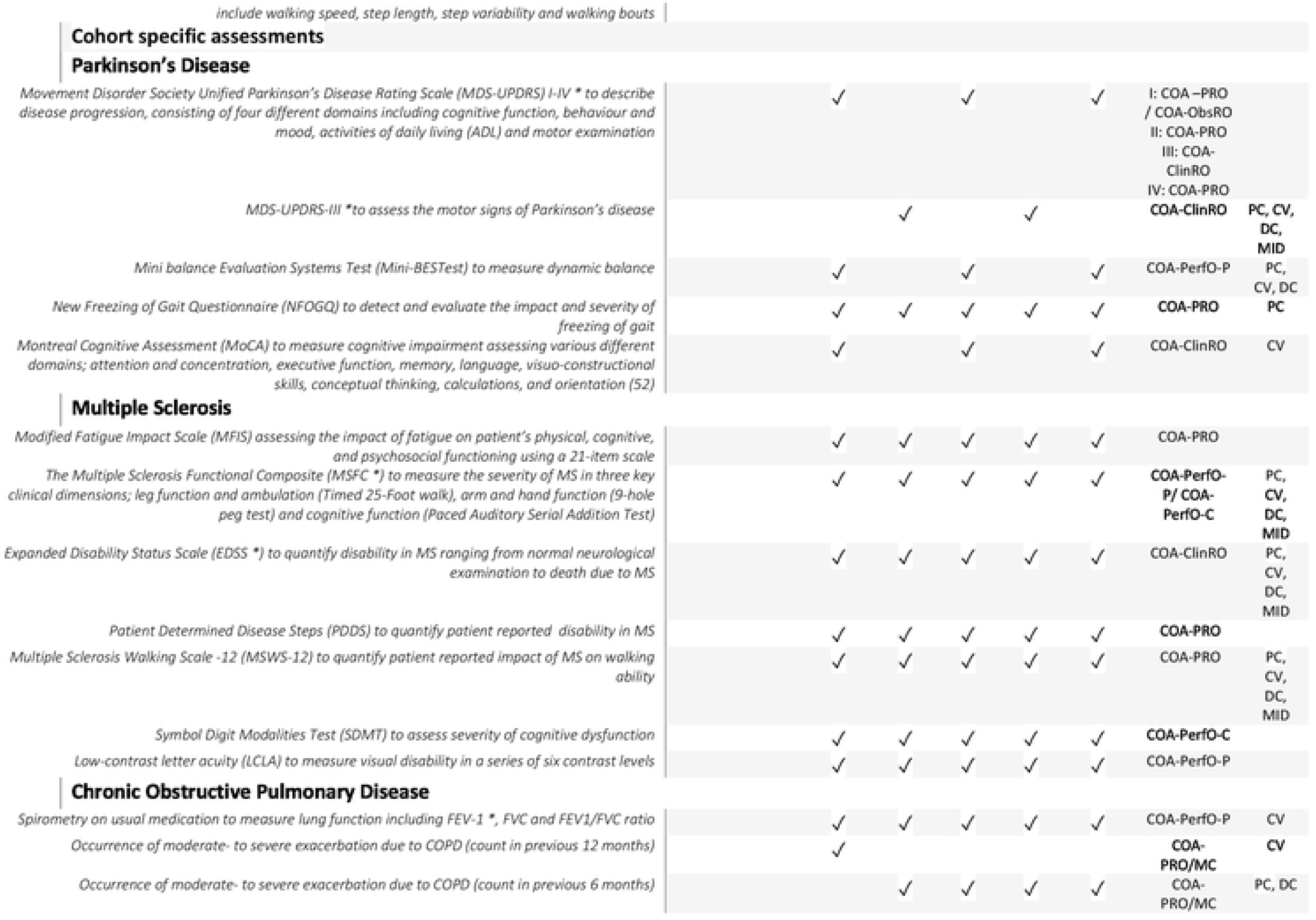
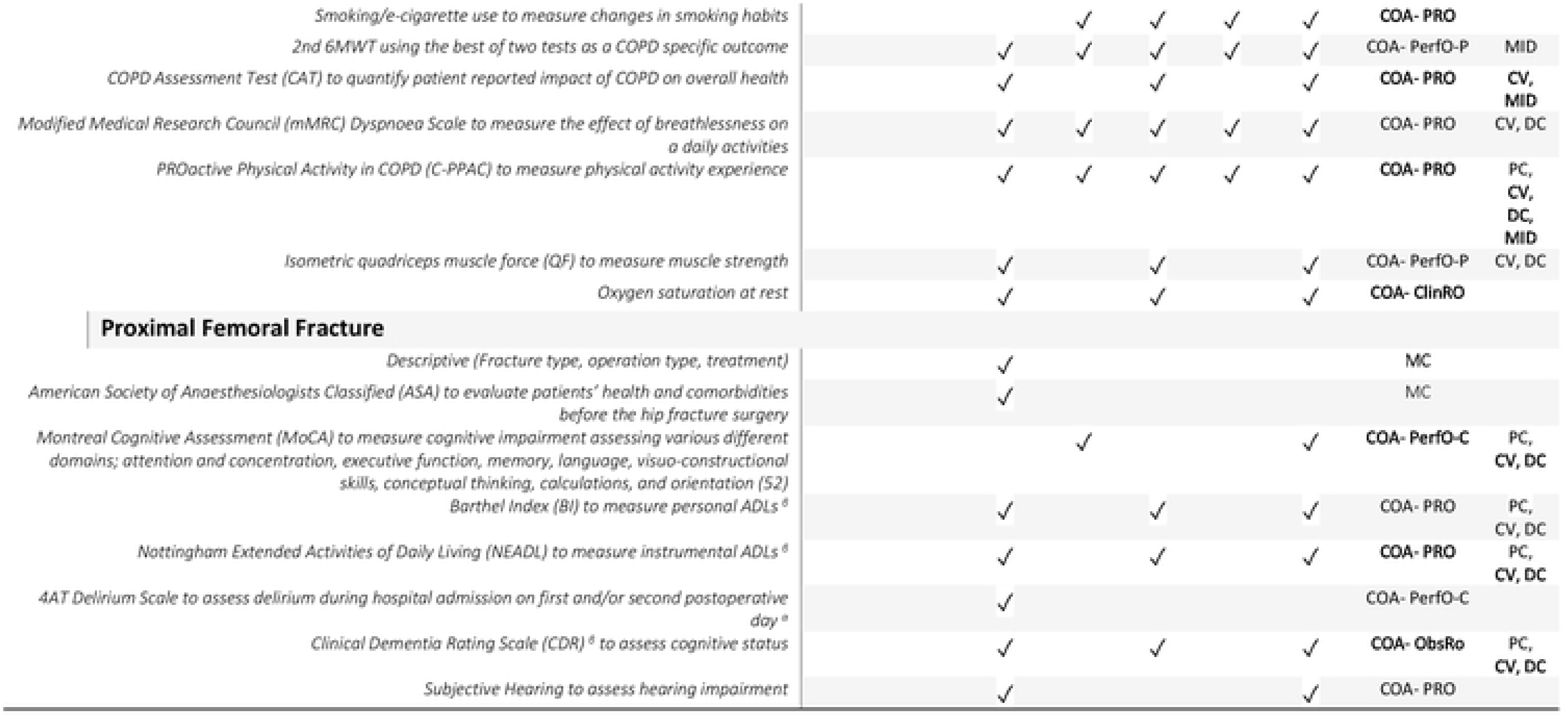
List of assessments and outcomes collected during screening, baseline assessment and every six months. Legend: T1, Screening/Baseline; T2, 6 month assessment; T3, 12 month assessment; T4, 18 month assessment; T5, 24 month assessment; *, indicates key (primary) cohort specific outcome measure; SPPB, short physical performance battery – PFF key primary cohort specific outcome measure; † falls and fracture data are collected retrospectively, 12 month retrospective at T1 and 6 month retrospective at T2-T5; β, pre-fracture status is measured at T1, current status is measured at T3 and T5; α, only applicable to acute patients; Outcome type, type of outcome measure in accordance with FDA terminology; COA, clinical outcome measure – describes or reflects how a patient feels, functions, or survives; PRO, Patient-reported outcome; ObsRO – Observer-reported outcome; ClinRO, Clinician-reported outcome; PerfO, Performance-based outcome; PerfO-P, Performance-based outcome physical measure; PerfO-C, Performance-based outcome cognitive/mental measure; Construct, validation construct assessed; PC, predictive capacity; CV, construct validity; DC, detect change over 24 months; MID, Minimum Important Difference; MC, medical chart;

### Study design and setting

Mobilise-D CVS is a longitudinal, observational cohort study, funded by the EU Innovative Medicine Initiative (IMI), which will include 2400 participants from four disease cohorts recruiting from 17 clinical sites across ten countries (Belgium, France, Germany, Greece, Israel, Italy, Norway, Spain, Switzerland, United Kingdom).

### Study procedure and study flow

#### Recruitment and screening

Participants are identified through research registries, out-patient and in-patient services. Potentially eligible and interested participants are invited to a screening appointment during which informed consent is obtained and eligibility is confirmed. (Informed consent forms available within the appendices). Rates of eligibility are monitored. Details of the recruitment process are shown in Figure . Each participant will be followed-up every 6 months for a total of 24 months Figure); screening and baseline assessment (T1) must be completed at the respective clinical site; T2 to T5 should also be completed at site, but can be completed at the participants’ home under exceptional circumstances to minimise drop-out and loss to follow-up.

Participants’ flow through the study is documented and presented in a Consolidated Standards of Reporting Trials flow diagram (41). Reasons for dropout are recorded and data collected up to the time of withdrawal will be included in analyses unless the participant withdraws their consent.

**Figure 2:**
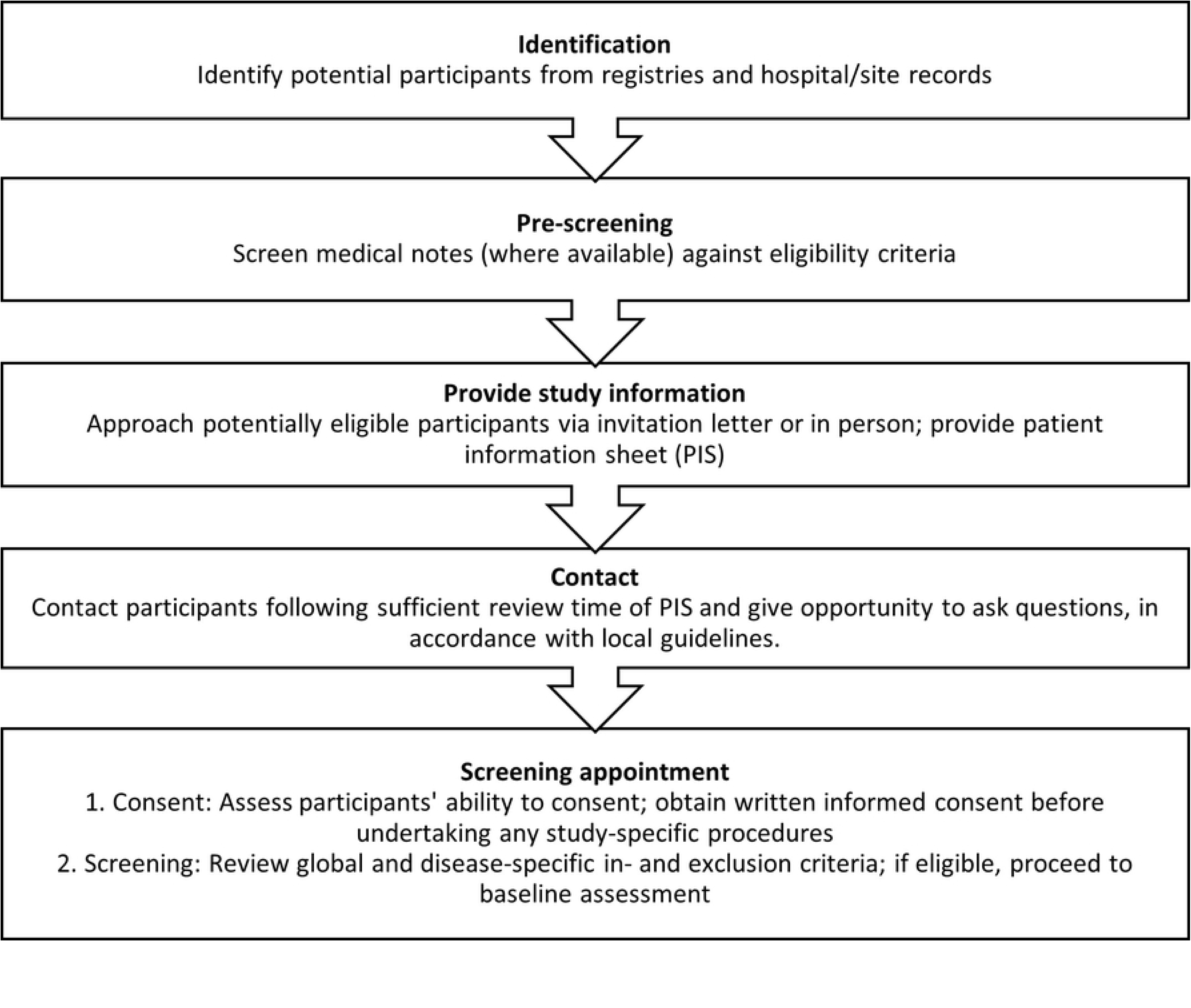
Flow chart to illustrate full recruitment process.

**Figure 3:**
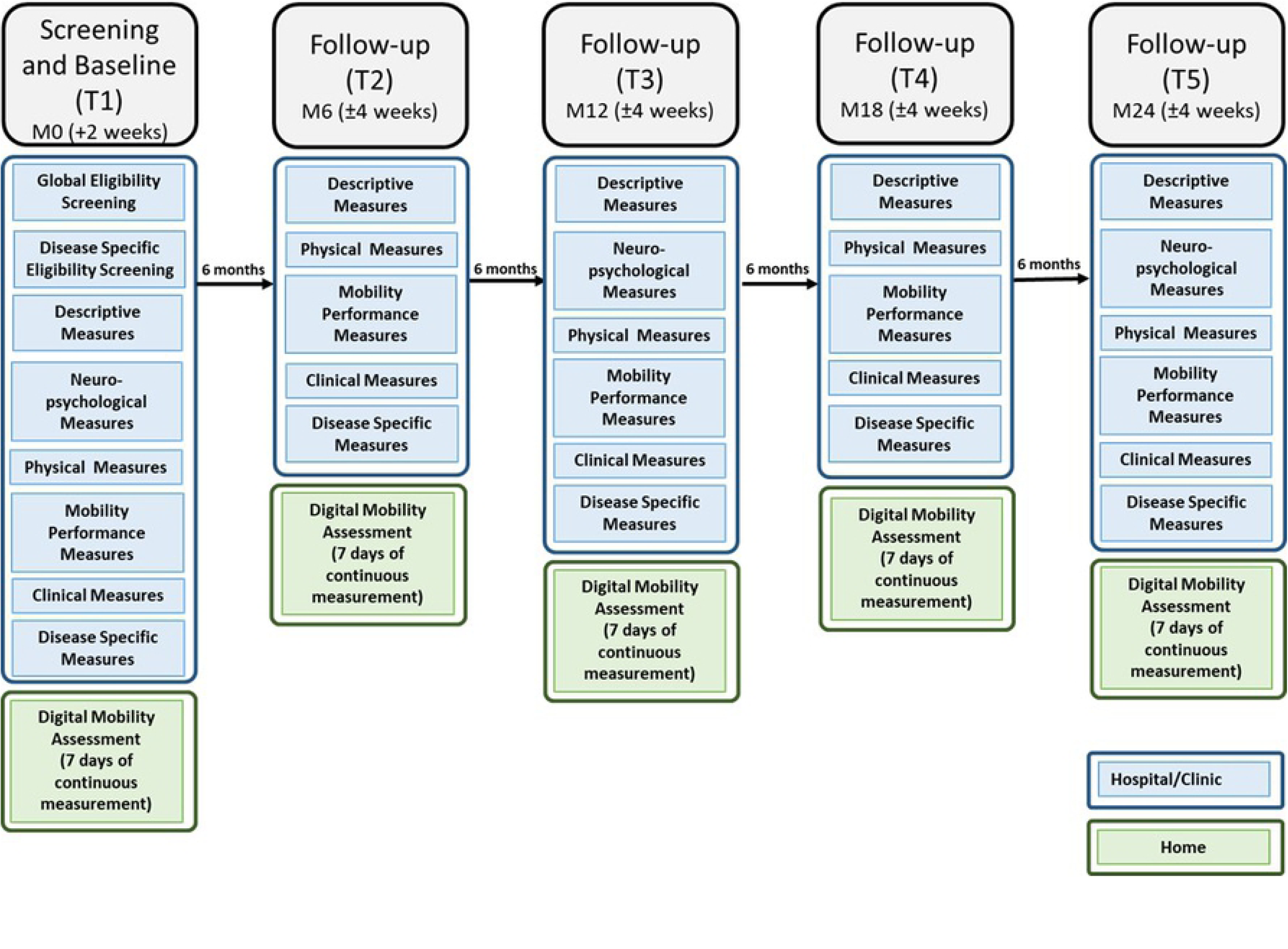
Study flow.

### Participants

The four disease cohorts studied within Mobilise-D CVS are Parkinson’s Disease (PD), Multiple Sclerosis (MS), Chronic Obstructive Pulmonary Disease (COPD) and Proximal Femur Fracture (PFF). Participants have to meet the core eligibility criteria as well as their respective cohort eligibility criteria. The full inclusion and exclusion criteria are listed in Table 1.

**Table 1:** Inclusion and Exclusion Criteria.

| Cohort | Inclusion Criteria | Exclusion Criteria |
| --- | --- | --- |
| **All** | • Able to walk 4 meters independently with or without walking aids<br>• Anticipated availability for repeated study visits over 24 months<br>• Ability to consent and comply with any study specific procedures.<br>• Willingness to wear a wearable sensor for mobility monitoring<br>• Able to read and write in first language in the respective country | • Occurrence of any of the following within 3 months prior to informed consent: myocardial infarction, hospitalization for unstable angina, stroke, coronary artery bypass graft (CABG), percutaneous coronary intervention (PCI), implantation of a cardiac resynchronization therapy device (CRTD), active treatment for cancer or other malignant disease, uncontrolled congestive heart disease (NYHA class >3), acute psychosis or major psychiatric disorders or continued substance abuse |
| **PD** | • Aged 18 or over<br>• Patients with the clinical diagnosis of PD according to the recent criteria of the Movement Disorder Society<sup>2</sup><br>• Hoehn & Yahr stage I-III | • History consistent with Dementia with Lewy Bodies (DLB), atypical parkinsonian syndromes (including multiple system atrophy or progressive supranuclear palsy, diagnosed according to accepted criteria)<br>• Repeated strokes or stepwise progression of symptoms, leading to a diagnosis of 'vascular parkinsonism'<br>• Drug-induced Parkinsonism |
| **MS** | • Aged 18 or over<br>• A diagnosis of MS based on the revised McDonald's criteria<br>• EDSS score of 3.0-6.5<br>• Clinical evidence of disability worsening over the previous two years | • Clinical relapse within 30 days prior to screening and baseline. |
| **COPD** | • Aged 18 or over<br>• Diagnosis of COPD (post-bronchodilator forced expiratory volume in the first second (FEV<sub>1</sub>) to forced vital capacity (FVC) ratio <0.70<br>• Clinical stability, defined as at least 4 weeks after the onset of the last exacerbation<br>• Current or ex-smokers with a smoking history equivalent to at least 10 pack years (1 pack year = 20 cigarettes smoked per day for 1 year) | • Having undergone major lung surgery (e.g. lung transplant)<br>• Current diagnosis of lung cancer<br>• Primary respiratory diseases other than COPD<br>• Substantial limitations in mobility due to factors other than COPD<br>• Lung volume reduction within 6 months before inclusion |
| **PFF** | • Aged 45 or over<br>• Surgical treatment (fixation or arthroplasty) for a low-energy fracture of the proximal femur (ICD-10 diagnosis S72.0, S72.1, S72.2) as diagnosed on X-rays of the hip and pelvis. Between 3 days and 52 weeks post-surgery | • Not able to walk before treatment of hip fracture |

The disease cohorts have been chosen because they represent different classes of mobility problems relating to low physical activity, different gait disturbances, and frailty, each affecting large groups of European citizens over substantial periods of time. The disease cohorts include a broad and heterogeneous range of subject characteristics with varying chronic care needs, and represent different trajectories of disability.

### Cohort descriptions

In the PD cohort, participants with mild to moderate disease state will be included (Hoehn & Yahr stage 1-3), which is of interest due to: 1) PD being a progressive disease, so the evaluation of a mobility endpoint over the course of two years can provide insight into disease trajectories; 2) PD being a heterogeneous disease, so the assessment of mobility endpoints in this disease can give relevant insight into the sensitivity to change of this endpoint across a broad range of mobility patterns; 3) improving our understanding of the association of mobility disability with falls and the influence of PD-specific symptoms on mobility in particular; and 4) PD being associated with characteristic gait disorders that can validate the Mobilise-D gait algorithm also for “challenging” gait deficits.

In the COPD cohort, patients with different COPD severity will be included, from those with mild disease and little burden of disease to those with very severe disease and significant burden and mobility impact. This represents the typical sample of patients recruited in clinical trials investigating pharmacologic and non-pharmacologic interventions. Exacerbations are events that punctuate the disease progression in COPD and their occurrence marks moments of acute deterioration and are often followed by only partial recovery. In the COPD cohort particular attention will be given to these events and how they interact with mobility in particular mobility decline. Mobilise-D will allow the study of mobility worsening over time in relation to other relevant clinical outcomes. In addition, the collection of treatment data will allow identification of patients undergoing pulmonary rehabilitation programs which offers insight in mobility improvement trajectories.

The MS population will be based on the revised McDonald’s criteria, with mild to moderate disability., Patients will be recruited covering a wide spectrum of overall disability and varying severity of walking impairment, with Expanded Disability Status Scale from 3 - fully ambulatory but with mild to moderate disability in other functional systems - to 6.5 –indicating reduced walking distance and requiring walking aids to walk 20 meters without resting. They will also have had disability progression over the previous two years, to enrich the sample with patients who are likely to progress over the next two years. Patients with a recent relapse (30 days before screening) will be excluded in order to have a reliable mobility measure not impacted by the recovery from a relapse.

The PFF cohort will be recruited either during the acute postoperative or subacute phase (max. 52 weeks) following sustaining a hip fracture. All PFF participants are community dwelling at point of enrolment; care home residents will not be included due to their increased burden of comorbidities likely to bias mobility results. Enrolling acute PFF patients allows monitoring of mobility during the recovery phase over the first 12 months providing unprecedented analysis of mobility trajectories over a long period of time in a disease cohort where mobility impairment is the direct consequence of a major trauma. Enrolling subacute PFF participants will allow a longer follow-up of mobility to focus on functional decline in a population with a high prevalence of frailty and sarcopenia. The aim is to study a mixed population of older community-dwelling adults to capture those who received for informal and formal care as well as participants who were independent prior to sustaining a hip fracture.

### Ethics and regulatory approval

The Newcastle upon Tyne Hospitals NHS Foundation Trust (NuTH) is the sponsor for the entire study. NuTH is responsible for ensuring appropriate regulatory and ethical approvals are in place at all participating sites. A Site Agreement between the sponsor and each site is required. Local sites teams are responsible for submitting the study to their local ethics committee for approval. Sites must not start recruitment until the sponsor has issued the regulatory Green Light for them to do so. This involves confirmation that the site has appropriate staffing, resources and documentation in place to undertake the study. The sponsor is also responsible for approving all study amendments, and for reviewing protocol deviations and Serious Adverse Events.

### Study registration

The study was registered at the ISRCTN registry on 12/10/2020 titled Clinical validation of a mobility monitor to measure and predict health outcomes (ISRCTN Number: 12051706).

### Outcome measures

In order to meet the objectives of the CVS, digital mobility outcome measures as well as general and disease specific outcome measures (constructs) will be assessed.

### Digital mobility measures

Daily-living mobility performance and capacity will be assessed directly using wearable sensor devices. First, participants will wear the McRoberts MoveTest (McRoberts B.V., The Hague, The Netherlands) to ‘instrument’ and objectively quantify physical assessments. Following the agnostic device approach proposed in the technical validation study (36), two different but metrologically equivalent devices will be adopted for the daily living monitoring. Accordingly, either the McRoberts MoveMonitor+ or the Axivity AX6 (Axivity Ltd, Newcastle Upon Tyne, UK) will be used to record free-living activity data over a period of at least seven consecutive days in an unsupervised condition after completion of the on-site assessment. Patients will be followed up using the same device at each time point. The MoveMonitor+ will be worn in a belt that is secured around the waist of the patients, while the AX6 will be directly secured to the skin using an adhesive fixation method. Mobilise-D walking-related DMOs include, among others, real world walking speed, quantitative measures of the total number of walking bouts, the percent of time spent walking which reflects the total walking in relation to the overall walking and non-walking activity, stride/step duration, median walking bout duration, median number of steps, and median cadence per bout. Quality-related sensor-derived measures include frequency-derived measures that reflect variability of the gait pattern such as swing and stance phase duration, variability and asymmetry of step time, stride time, swing time, stance time and velocity stride length which quantity gait rhythmicity and consistency (36). This further aligns with the categories of DMOs identified recently which are pace, volume, rhythm, phases, variability, base of support and asymmetry of gait (35).

### Clinically relevant outcome measures (constructs)

A wide range of core and cohort specific outcome measures will be collected, spanning patient-reported outcomes (PRO), observer-reported outcomes (ObsRO), clinician-reported outcomes (ClinRO), performance-based outcome physical measures (PerfO-P), and performance-based outcome cognitive/mental measures (PerfO-C). The assessments will be completed according to the assessment schedule (**Error! Reference source not found.**), with all outcome measures detailed in Figure 1.

### Primary outcome measures (Global and cohort-specific)

There is one global primary outcome measure and four cohort specific primary outcome measures. The Late-Life Function and Disability Instrument (LLFDI) (42, 43) represents the core primary outcome measure. The LLFDI was developed as a comprehensive questionnaire assessing function and disability for use in community-dwelling older adults. It contains items that represent functional limitations in performing discrete physical tasks encountered in daily routines and disability related to participation in major life tasks and social roles within a typical sociocultural and physical environment. The LLFDI assesses function in 32 physical activities in three dimensions (upper extremity, basic lower extremity, and advanced lower extremity) and disability in 16 major life tasks.

The cohort specific primary outcome measures are fall frequency during 24-month follow-up in the PD and MS cohorts, occurrence of moderate to severe COPD exacerbations during the first 12-months follow-up in the COPD cohort and admission to a care home at six-months follow-up within the PFF cohort.

### Secondary outcome measures

Secondary outcome measures encompass sociodemographic (descriptive data) and clinical outcome measures including physical measures, mobility performance, and neuropsychological measures to describe how participants feel, function, or survive (Figure 1).

Primary and secondary outcomes will be used as the constructs against which the DMO’s predictive ability, construct validity, estimates of the Minimum Important Difference, and ability to detect change over 24 months will be assessed. Figure 1 includes details of all assessments and outcome and their respective validation use.

The secondary outcome measures of special interest for each cohort are as follows:

The Movement Disorder Society Unified Parkinson’s Disease Rating Scale (MDS-UPDRS) (44) will be used to assess predictive capacity, construct validity, detect change over 24 months as well as determine the MID amongst the PD cohort. The MDS-UPDRS describes disease progression, and it is separated into four different domains including cognitive function, behaviour and mood, activities of daily living (ADL), and motor examination.

The Expanded Disability Status Scale (EDSS) (45) will be used to assess predictive capacity, construct validity, detect change over 24 months as well as determine the MID amongst in the MS cohort. The EDSS is an ordinal clinical rating scale to quantify disability in MS ranging from normal neurologic examination (0) to death due to MS (10).

Forced expiratory volume in one second (expressed as a percentage of predicted norm) (FEV-1) will be used to assess predictive capacity, construct validity, detect change over 24 months as well as determine the MID amongst for the COPD cohort. The spirometry test measuring FEV-1 is used to diagnose and stage COPD.

The Short Physical Performance Battery (SPPB) (46) will be used to assess predictive capacity, construct validity, detect change over 24 months as well as determine the MID amongst the PFF cohort. The SPPB assesses lower extremity function and mobility, consisting of a static balance task, a five-repetition chair-rise test, and a 4-meter walk test.

### Diaries

MS and PD participants will be asked to complete a falls diary and return these on a monthly basis recording the relevant events (exacerbation, change of medication, falls) as and if they occur during each month. COPD participants will be asked to complete exacerbation diaries including medication change, hospitalisation, unplanned doctor’s visits, and falls and return diaries at subsequent study visits. Other measures

### Environmental factors

Given the fact that the CVS will not be carried out under controlled laboratory settings, meteorological variables (such as daily maximum temperature, precipitation, snowfall, and mean wind speed, among others) will be collected to characterise the participants’ weather condition exposure and assess the effect of these environmental factors on the digital mobility outcomes.

### Patient and Public Involvement and Engagement (PPIE)

Prior to the design of the CVS, patients’ opinions regarding acceptability of wearable devices to measure gait and physical activity were collected and explored. Patients were involved in the design of the study protocol, including the consideration of COVID-19 protocols and risks, and in the review of patient-facing documents to ensure readability and understanding. Throughout the CVS, a dedicated Patient and Public Advisory Group (PPAG) will advise on key topics such as the identification of meaningful mobility outcomes and patient needs and concerns regarding digital technology. Furthermore, the PPAG will co-design and comment on research plans, protocols and materials, and assist in the interpretation of results and their dissemination.

### Management, Safety Monitoring, and Steering Committee

Formal oversight of the Mobilise-D CVS will be undertaken by the Study Management Group (SMG) and the independent clinical Study Steering Committee (SSC). Informal oversight of the Mobilise-D CVS will be undertaken by the Assessor Support Group (ASG). All committees will run for the study duration.

#### Study Management Group

The SMG will be responsible for the day-to-day management of the study. The group includes the key individuals responsible for undertaking the study. The SMG will monitor the study progress, ensure protocol adherence, and take appropriate action to safeguard participants and the quality of the study. Data issues and safety will be a priority for the SMG. This is a low risk observational study that does not require a specific Data Monitoring Committee.

#### Study Steering Committee

The SSC will provide overall supervision of the study. The SSC has an independent chair and majority independent representation including patient representatives. The SSC will monitor the study progress and is responsible for making top-level decisions. The SSC carries the responsibility for deciding whether the study should be stopped on grounds of safety or efficacy.

#### Assessor Support Group

The ASG will be responsible for the day-to-day running of the study and to provide general support to study assessors. The ASG is managed by the study coordinators. Individual cohort leads are responsible for providing cohort specific oversight and support.

### Statistical analysis

#### Sample size

Sample size was calculated separately for each cohort according to its main disease-specific primary outcome, as well as with 24-months changes in LLFDI score as global primary outcome. The estimated sample size aiming for a sample that allowed the disease-specific objectives as well as the global objectives to be met.

#### Cohort specific sample size calculations

Sample size calculation for each disease cohort showed that 600 participants will be required in each cohort in order to achieve a statistically significant outcome of the respective hypothesis critical value with a power of 90% and an alpha error of 0.05. Details of the hypotheses and the assumption underpinning these calculations are summarised in Table 2.

**Table 2:**
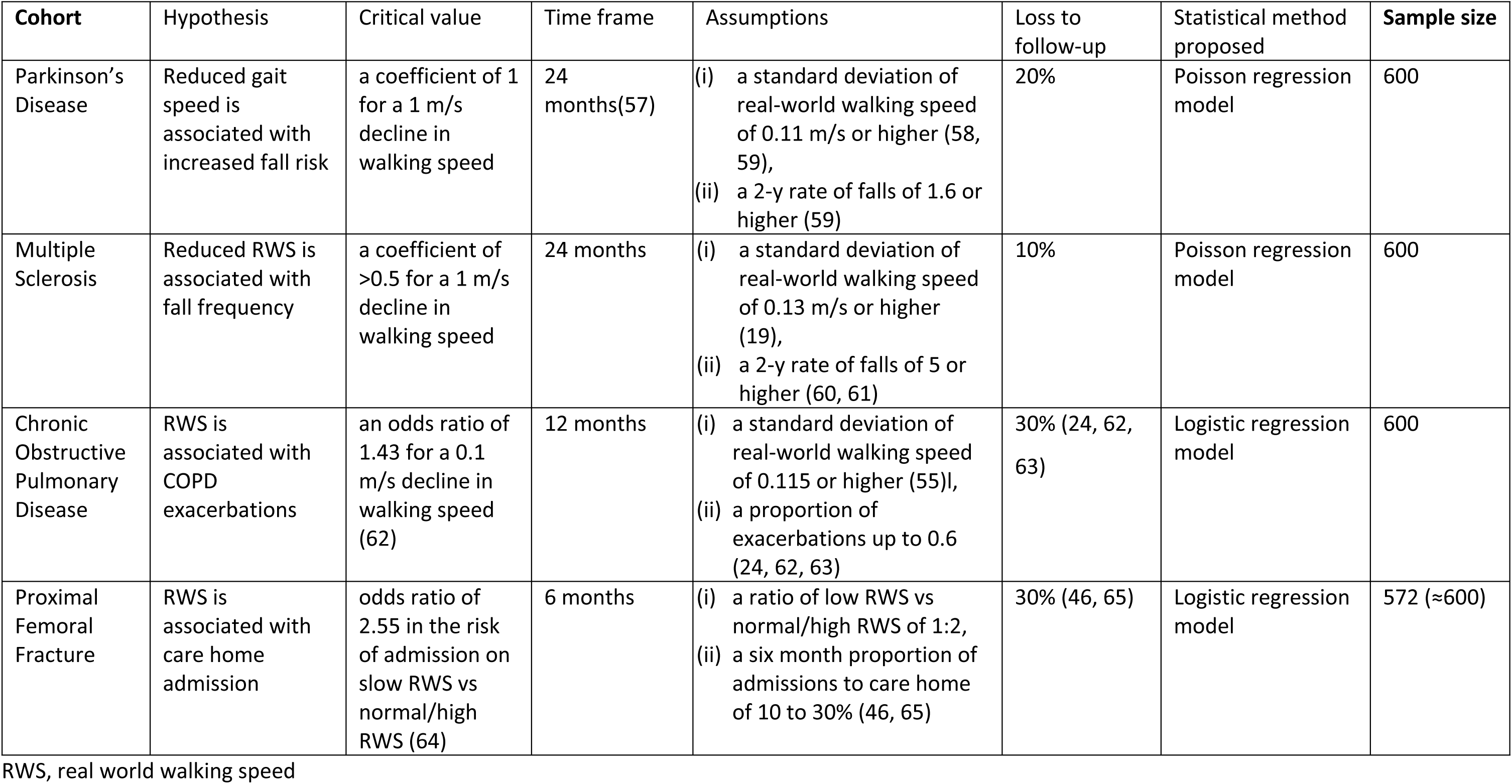
Cohort specific sample size calculation.

#### Global sample size calculations

Sample size calculation for the global primary outcome is based on the hypothesis that baseline real-world walking speed is associated with changes in LLFDI score (0–100) within 24 months. Using (i) a minimal detectable change of 95% confidence of 3.41 in LLFDI functional component (53) representing a clinically relevant intervention effect (54), (ii) a standard deviation of the mean change in LLFDI functional component of 30.1 points (41), (iii) a standard deviation of 0.29 m/s for real-world walking speed (55), (iv) a clinically relevant and measurable difference of 0.1 m/s in real-world walking speed between two patients (56), and (v) based on a linear regression analysis a sample size of 67 subjects would allow identifying a statistically significant change, with a power of 80% and an alpha error of 0.05. Including an expected drop-out rate of 10-30% the final sample size would be n=81. The available sample size of 2400 patients, derived from the sum of sample sizes required for the evaluation of the abovementioned cohort-specific outcomes provides sufficient power to test the general LLFDI hypothesis.

The proposed statistical method will be amended upon data completion and prior analysis should the data distribution require a more appropriate methodology.

### Analysis plan

The statistical analysis will follow a pre-specified step-wise procedure. We will elaborate a Statistical Analysis Plan including the definition of analysis sets, details on data edition (including derivation of new variables), handling of missing data and statistical analysis (including prioritisation of outcomes).

Descriptive analysis of main characteristics of patients, including detailed description of COAs and DMOs, will be done by number and percentage for categorical variables, mean and standard deviation for continuous variables with normal distribution, and median and percentiles 25th-75th for continuous variables with non-normal distribution.

We will test construct validity (convergent and known-groups). For convergent validity, we will test the correlation (Pearson or Spearman, depending on variables distribution) between DMOs and related constructs. A table of expected correlations for each DMO-construct combination will be built based on existing literature. For known-group validity, we will use one-way ANOVA test and pairwise comparisons of means between groups a priori expected to have differences in DMOs values. To test predictive capacity of DMOs against the disease-specific and global outcomes we will estimate the association between baseline DMO levels and each outcome using multivariable regression models (specific models depending on outcome distribution) adjusting for confounders. Non-linear associations will be tested using generalised additive models and appropriate transformation of variables will be done consequently. Secondary analyses will include use of DMO changes over time as predictors, and adjusting for baseline levels as sensitivity analysis. The ability of each DMOs to predict disease-specific and global outcomes will be compared to that of traditional predictors and predictive scores.

To quantify the ability to detect change of DMOs, we will calculate the change (between baseline and different follow-up periods) and the standardised response mean (SRM) in (1) groups defined by the self- reported change in mobility, (2) groups defined according to having had a clinically relevant event (e.g., an exacerbation in COPD, a fall in Parkinson) during follow-up, and (3) groups defined according to clinically relevant changes in anchors.

We will establish the MID by triangulation using anchor- and distribution-based estimates. To describe the real-world walking behaviour we will extract walking behaviour metrics during the course of the day and week and model them using advanced statistical analysis such as cumulative probability function, detrended fluctuation analysis or entropy analysis.

### Data Management

All study data will be transferred to a centrally managed data management platform. **Error! Reference source not found.**The Mobilise-D data management platform has been created on e-SC, an open-source cloud-based platform designed to provide secure ingestion, storage, sharing, and analysis capability for scientific studies (www.esciencecentral.co.uk) (66). Participant data is collected in a coded, de-identified manner, using electronic data capture as a default option. Data is either entered into the Mobilise-D data management platform directly or via third-party platforms (e.g., McRoberts or ERT). The proposed data flow model is illustrated in **Error! Reference source not found.**Figure . Where electronic data capture is not feasible, paper case report forms are used (e.g., falls diaries). Data from paper forms is entered into web forms on e-Science Central (e-SC), and the signed and dated paper forms will be scanned and uploaded to the platform. The Mobilise-D e-SC platform has been implemented using Amazon Web Services located within the European Union.

Direct access will be granted to authorised representatives from the Sponsor, host institution, and the regulatory authorities to permit study-related monitoring, audits, and inspections- in line with participant consent. Clinical sites will be responsible for archiving all study documents for a period of time that is in keeping with institutional or national guidelines that pertain to that site.

**Figure 4:**
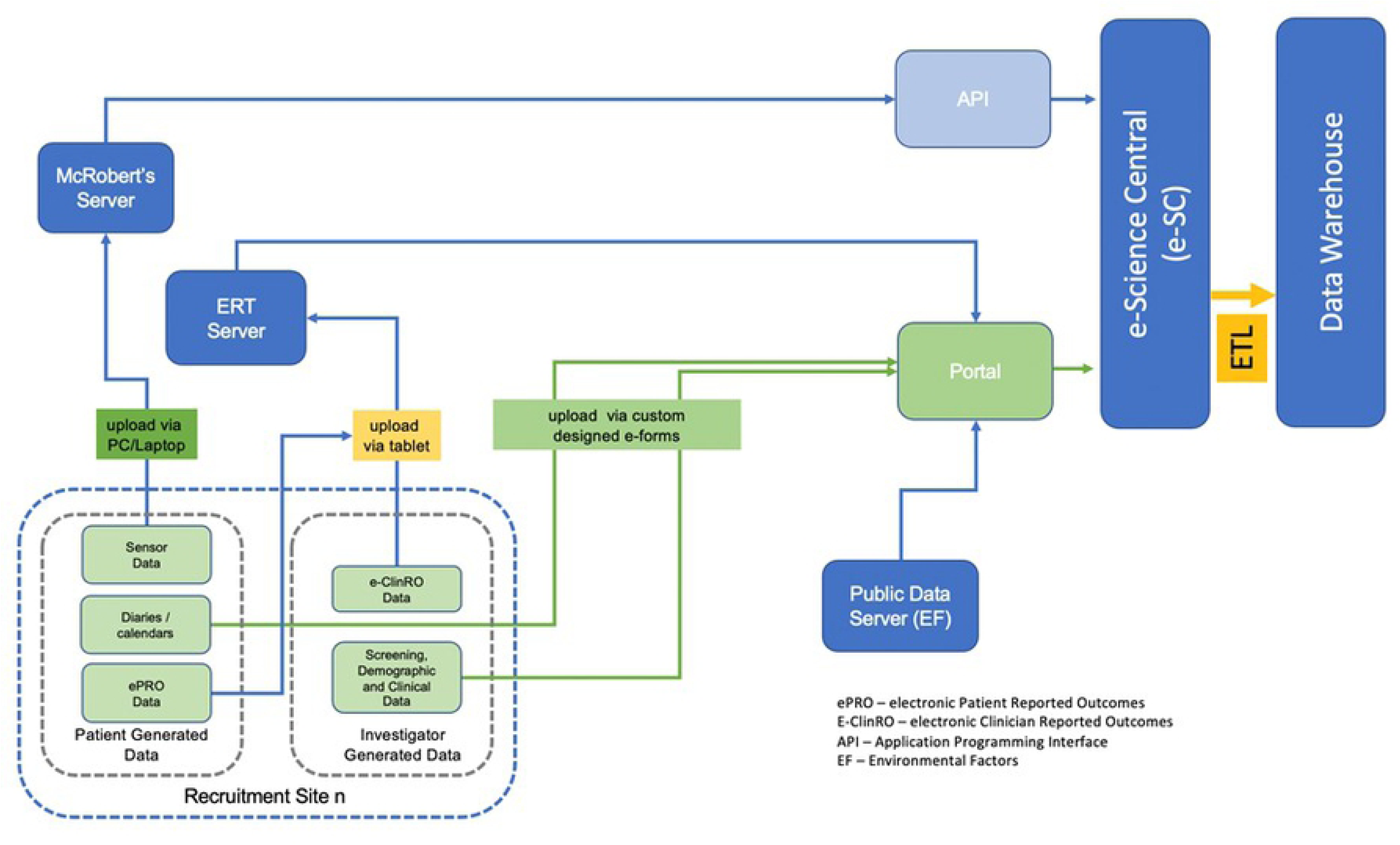
Proposed Data Flow.

### Data monitoring

Central monitoring of recruitment and data quality will be undertaken by the Mobilise-D Data Manager, as documented in Study Monitoring Plan. Participant enrolment will be regularly reviewed and compared against expectations for site and disease cohort. Where recruitment falls below a pre-defined percentage, mitigation plans identified by sites will be actioned. All data quality monitoring will be performed centrally, for example on data within study databases, uploaded documentation or by self-assessment checklists at site. Any data queries identified through central monitoring will be sent to each site in form of a monthly report. On-site visits will only be triggered if serious issues are identified at a site level. There is no non-serious adverse event reporting planned for this study. However, any Serious Adverse Events (SAEs) that occur during the study visits must be reported to the sponsor and will be recorded on a central SAE log.

### Staff training

A comprehensive training package has been developed, including a core manual to support delivery of the protocol robustly and reliably. In addition, four cohort-specific manuals are prepared in which the cohort-specific assessments are addressed in the same manner as in the core manual, i.e., administration, scoring, and troubleshooting of assessments are described. A third manual describes the exact procedures of instrumented physical assessments as well as the 7-day mobility assessment. A series of eleven webinar sessions has been designed and will be held on a weekly basis to cover all relevant aspects of the manual, including additional parts concerning data entry platforms, data management procedures, and practical training sessions. All study personnel assigned to the clinical work package are invited to participate in the webinars, i.e., assessors, site coordinators, and site lead across the project. Several background topics are addressed, for which expert members of the Mobilise-D consortium are invited as speakers. An additional webinar per cohort will be held to address the specific issues related to the four different cohorts. To ensure that all current and future assessors receive the same training and information all webinars are recorded and stored online. Alongside the webinars, nine videos have been produced to describe measurement procedures of several assessments.

### Covid mitigation

As Europe is being severely affected by the COVID-19 pandemic, there have been and will be major impacts in the execution of the study. From a study procedure perspective, all activities have been transferred to a virtual/remote working environment including all assessor training being held via webinars and all sites monitored remotely. Two main impacts may be a slower recruitment rate and an increased drop-out rate. The reporting and monitoring of enrolment of new patients will be tracked through weekly meetings. Contingency plans for all cohorts are in place to expand recruitment sites (e.g., other clinics or hospitals within the geographical area) and recruitment pathways (e.g., via registries, clinics, other forms of advertisement) within each country, and also to commence recruitment of cohorts at sites primarily recruiting a different cohort.

### Dissemination policy

We will seek to publish all results from this clinical validation study in open-access, peer-reviewed international journals, and disseminate findings at scientific and non-scientific conferences and events. Main results will also be shared on the project website and spread to various stakeholders. Authorship eligibility will follow the International Committee of Medical Journal Editors.

The algorithms and procedures which result from the consortium efforts will be made publicly available at the end of the Mobilise-D project, enhancing future research and development activities.

## Discussion

Digital methods that consistently and accurately measure the extent and nature of mobility are now within reach. Once implemented, they will become a benchmark for new interventions, improving patient outcomes in a manner that will extend well beyond the cohorts studied in Mobilise-D.

The assessment battery including feasibility advice will be made available during the final year of the project to provide advice for future trials, including observational studies and large-scale epidemiological studies to optimize the combination of clinical outcome assessments (COAs) and Digital Mobility Outcomes (DMOs) that are useful as digital biomarkers for monitoring, evaluation, prediction, stratification, or safety evaluation. Further, we will report on the feasibility of the assessment battery in its entirety after the baseline assessments have been completed (estimated mid 2022) given that the current protocol is very lengthy due to capturing participants of various ages, across four cohorts, with different disease activity states and on different trajectories of decline. Floor and ceiling effects will be reported and considered in the revisions as well as recommendations on appropriateness of the outcome measures used within Mobilise-D.

Due to the heterogeneity of participants in the planned study sample, a consensus and compromise on parameters collected during assessment had to be reached. This has resulted in well-established and cohort specific assessments (e.g., 6-min walk test in COPD) to be undertaken in other cohorts where these have not been commonly used so far (e.g., PD and PFF). However, this cross-cohort assessment consensus is paramount to establish harmonized, generalizable, and comparable results valid across chronic disease types and activity states. Only this way can common cohort specific assessments be considered applicable across cohorts. The results of Mobilise-D will directly benefit drug development and monitoring, and establish the roadmap for clinical implementation of new, complementary tools to identify, stratify and monitor disability. The objective is to enable widespread access to optimal clinical mobility management through personalised healthcare facilitating cost-effectiveness and cost-utility appraisal. This meets the need recognized by the Committee for Medical Products for Human Use (CHMP) for monitoring real-world mobility and standardizing digital mobility assessment. Within Mobilise-D, we will be able link patients’ capacity of mobility and their actual performance in a real-world setting, which will provide meaningful, robust, harmonized, and objective measures for regulatory evaluation of disease interventions using a similar framework across different diseases.

Our low-cost technology has the promise to pervade clinical practice, with every healthcare centre able to offer inexpensive digital assessment systems, linked to a centralized system directly connected to healthcare databases. Our implementation of Mobilise-D will support advances in both therapy and care provision, with an opportunity for improved and more informed decision-making in assessing the efficacy of new interventions.

Mobilise-D will provide the first-ever systematic approach to mobility determination that is standardised, codified, and freely available to industry, extending patient measurement systems that have utility across all aspects of patient care, from research through intervention development to routine large scale patient care. Mobilise-D will develop and publish a new generation of clinically validated, open-access algorithms, associated reference data, and a defined battery of relevant digital mobility outcomes that will allow any patient to receive a consistent and accurate mobility assessment from simple, low-cost devices. It provides a model for future studies in the field of digital health assessment.

## Trial status

Staff training for the clinical validation study commenced in January 2021 and overall, each assessor received training during 12 webinars, which amounts to about 24 hours of training. Following ethical approval at sites and the establishment of all relevant management and steering committees, recruitment commenced in April 2021, with current recruitment expected to continue until 31 March 2023. As of March 2022, n=1,800 participants of the study sample have been recruited.

## Data Availability

No datasets were generated or analysed during the current study. All relevant data from this study will be made available upon study completion.

## Acknowledgements

Full membership of the Mobilise-D consortium is available on the website https://www.mobilise-d.eu/partners. ISGlobal acknowledges support from the Spanish Ministry of Science, Innovation and Universities through the “Centro de Excelencia Severo Ochoa 2019-2023” Program (CEX2018-000806-S), and support from the Generalitat de Catalunya through the CERCA Program. Heleen Demeyer is a post- doctoral fellow of the FWO Flanders. Heiko Gaßner is supported by the Fraunhofer Internal Programs under Grant No. Attract 044-602140 and 044-602150.

## Supporting information

S1 SPIRIT checklist

